# Genome-wide association study reveals a novel tuberculosis susceptibility locus in multiple East Asian and European populations

**DOI:** 10.1101/2024.03.14.24304327

**Authors:** Xuling Chang, Zheng Li, Phan Vuong Khac Thai, Dang Thi Minh Ha, Nguyen Thuy Thuong Thuong, Denise Wee, Alya Sufiyah Binte Mohamed Subhan, Matthew Silcocks, Cynthia Bin Eng Chee, Nguyen Thi Quynh Nhu, Chew-Kiat Heng, Yik Ying Teo, Amit Singal, Stefan H Oehlers, Jian-Min Yuan, Woon-Puay Koh, Maxine Caws, Chiea Chuen Khor, Rajkumar Dorajoo, Sarah J Dunstan

## Abstract

**Background:** Tuberculosis (TB) continues to be a leading cause of morbidity and mortality worldwide. Past genome-wide association studies (GWAS) have explored TB susceptibility across various ethnic groups, yet a significant portion of TB heritability remains unexplained.

**Methods:** We conducted GWAS in the Singapore Chinese and Vietnamese, followed by a comprehensive meta-analysis incorporating 4 independent East Asian datasets, resulting in a total of 11,841 cases and 197,373 population controls.

**Findings:** We identified a novel susceptibility locus for pulmonary TB (PTB) at 22q12.2 in East Asians [rs6006426, OR (95%Cl) =1.097(1.066, 1.130), *P_meta_*=3.31×10^-10^]. The association was further validated in Europeans [OR (95%Cl) =1.101(1.002, 1.211), *P*=0.046] and was strengthened in the combined meta-anlaysis including 12,736 PTB cases and 673,864 controls [OR (95%Cl) =1.098(1.068, 1.129), *P_meta_*=4.33×10^-11^]. rs6006426 affected *SF3A1* expression in various immune cells (*P* from 0.003 to 6.17×10^-18^) and *OSM* expression in monocytes post lipopolysaccharide stimulation (*P*=5.57×10^-4^). CRISPR-Cas9 edited zebrafish embryos with *osm* depletion resulted in decreased burden of *Mycobacterium marinum* (*M.marinum*) in infected embryos (*P*=0.047).

**Interpretation:** Our findings offer novel insights into the genetic factors underlying TB and reveals new avenues for understanding its etiology.

## Introduction

Tuberculosis (TB), an infectious disease caused by *Mycobacterium tuberculosis* (*Mtb*), poses a significant challenge to global health and is a major contributor to the rising global burden of antimicrobial resistance (1). The World Health Organisation (WHO) 2024 global TB report indicates a record high of 8.2 million new TB cases in 2023, exceeding the pre-COVID baseline of 7.1 million in 2019. TB predominantly affects individuals in low- and middle- income countries, with the highest burden observed in Southeast Asia, the Western Pacific, and African regions (1). Despite recent advances in treatment and diagnosis, the challenge to optimally disrupt transmission and ensure positive outcomes for all those infected remains. This enduring challenge is primarily attributed to several factors, such as reduced efficacy of the current vaccine, lengthy and complex multi-drug treatment, and the rising incidence of multidrug-resistant (MDR) and extensively drug-resistant (XDR) TB (2). Deeper understanding of TB etiology is crucial for identifying those at high risk and for development of effective vaccines and targeted treatments, including host-directed therapies, to control this global health threat.

TB susceptibility is influenced by a complex interplay of genetic and environmental factors (3, 4). Twin studies and mouse models demonstrate a strong host genetic influence on TB susceptibility (5, 6) with heritability estimated >50% (7). Since the advent of genome-wide association studies (GWAS) in 2005, significant strides in uncovering genetic contributions to diseases have been made, with the focus predominantly on non-communicable diseases in European (EUR) populations. Until September 2024, only 5,722 (0.84%) out of 681,210 associations recorded in the GWAS catalog (8), are infectious disease related. Additionally, the GWAS diversity monitor shows that 94.49% of GWAS participants are European (9). Therefore, comparatively, large-scale human TB GWAS has been neglected due to the disease being over-represented in resource-limited countries, particularly within populations that are under-represented in human genomics research. Despite this, research investigating TB susceptibility via GWAS has been conducted in populations from Africa (10, 11), Russia (12), Iceland (13) and Asia (14–16). However, these studies have not fully accounted for the genetic risk and have shown considerable location or ethnicity specific genetic associations, with minimal replication across populations (12, 13, 17).

Here we performed GWAS and meta-analysis to identify pulmonary TB (PTB) associated genetic variants shared among East-Asian populations. We first evaluated PTB case-control datasets from Singapore and Vietnam. To increase statistical power and confidence, we incorporated public data from Han Chinese (15) and Japanese (16), into the meta-analysis. We also evaluated the transferability of the association in a combined European dataset, from the UK biobank (UKB) and FinnGen (16). We identified a novel locus at chromosome 22q12.2 and prioritised *SF3A1* and *OSM* as candidate genes underlying the association by querying published TB transcriptomic and proteomic data alongside expression quantitative trait loci (eQTL) from the eQTL Catalogue (https://www.ebi.ac.uk/eqtl/) (18). Functional assessment of *osm* in the zebrafish-*Mycobacterium marinum* (*M.marinum*) infection model revealed a reduced burden of bacterial infection in the *osm* knockdown crispants, suggesting a protective role for *osm* suppression during infection.

## Results

### PTB Genome-Wide Association Study

We performed PTB GWAS using datasets from Singapore [the Singapore Chinese Health Study (SCHS)] (19) and Vietnam. After PCA-adjustment, no significant inflation was observed (Supplemental Figure 1), indicating that potential population stratification effects were well-controlled. No associations surpassing genome-wide significance in individual analyses (Supplemental Figure 1).

We next performed a meta-analysis incorporating all available PTB GWAS from East Asia. Published summary statistics from Han Chinese and Japanese (Biobank Japan) (15, 16) were merged with in-house GWAS data [4,581,617 overlapping single nucleotide polymorphism (SNPs)] resulting in a combined dataset of 11,841 PTB cases and 197,373 controls. The East- Asian meta-analysis indicated minimal inflation (λ=1.028) and identified two loci surpassing genome-wide significance (Figure 1a and 1b), including the known *HLA-DQB1* locus (16). The sentinel SNP in this meta-analysis was rs9274669 [OR (95%Cl) =1.160(1.121, 1.200), *P_meta_*=1.92×10^-17^, Supplemental Table 1], which is in weak linkage disequilibrium (LD) with the previously reported lead SNP rs140780894 (r^2^=0.114, JPT in 1000 Genome Project). The association of rs9274669 was primarily driven by data from Japan (*P*=7.84×10^-16^) and was significant in the Singaporean Chinese (*P*=0.002) while directional consistent in the Vietnamese and Han Chinese. We identified a novel locus at 22q12.2 [OR (95%Cl) =1.097(1.066, 1.130), *P_meta_*=3.31×10^-10^, Table 1, Figure 1a], which demonstrated significant associations across all included datasets (Table 1). The lead SNP, rs6006426, is an intergenic SNP 7,043 bp upstream of the transcription start site of the Oncostatin M gene (*OSM*) (Figure 1c), with a minor allele frequency (MAF) of 37%-47% in East Asian (EAS) populations according to the 1000 Genome Project (20).

**Figure 1.**
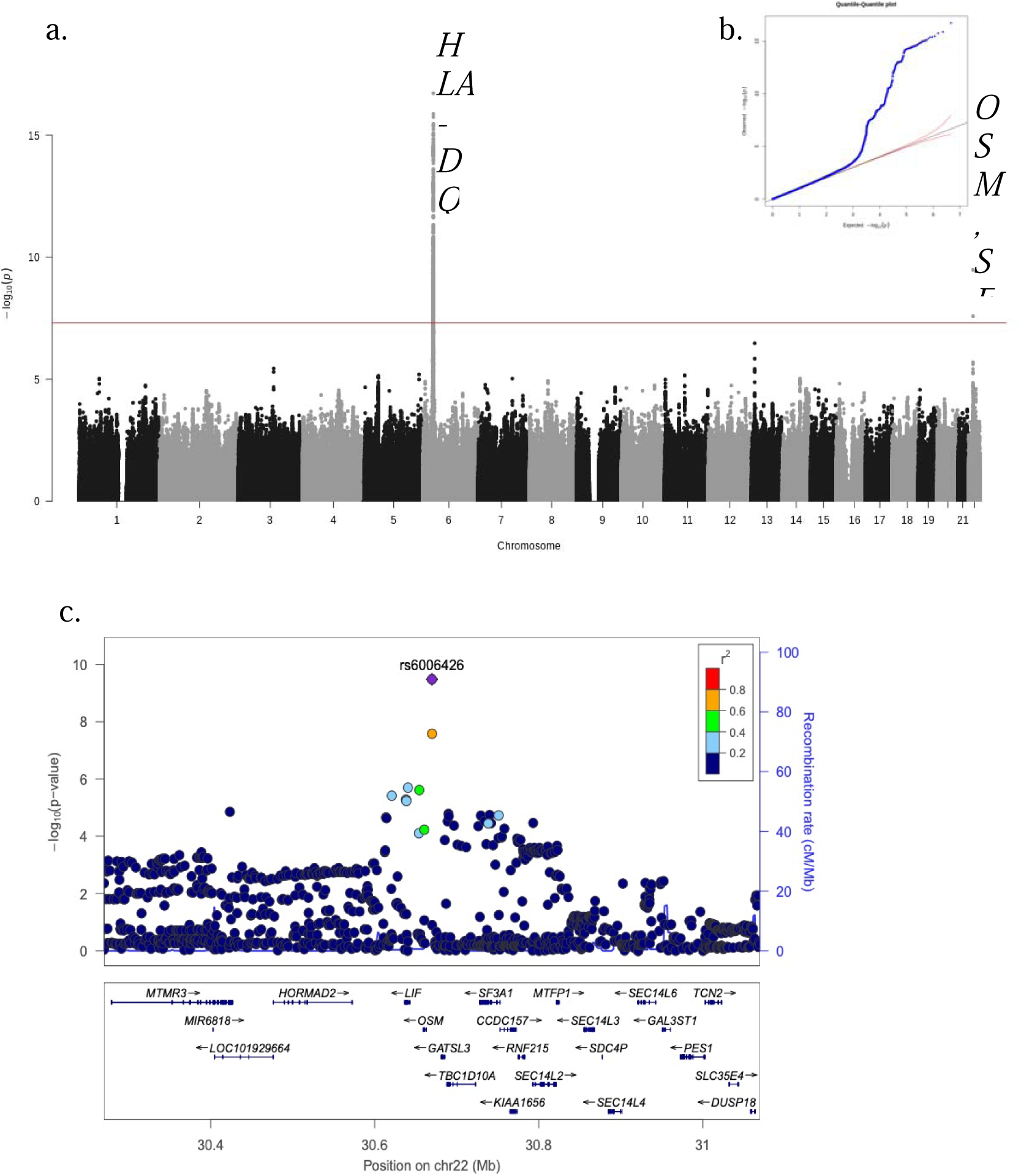
Meta-analysis for PTB in East Asia. **a** Manhattan plot of the meta-analysis of 4 Asian populations (11,841 cases/197,373 controls). Two hits at chromosome 6 and 22 were identified beyond the genome-wide significance threshold (*P*L<L5L×L10^-8^, red line) **b** QQ-plot of observed compared to expected *P*-values indicated minimal inflation (λL=L1.028). **c** Regional SNP associations at 22q12.2 in the meta-analysis of 4 Asian populations. Lead SNP indicated as purple diamonds. LD (r^2^) data of SNPs based on ASN panels of 1000Genome database. Plots plotted using LocusZoom (http://locuszoom.org/).

**Table 1.** Summary statistics of genome-wide associations identified in individual datasets and after meta-analysis for pulmonary tuberculosis.

|  | SNP | chromosome | position | Other allele | Effect allele | Sample size (cases/controls) | OR (95%CI) | *P* | *P*<sub>heterogeneity</sub> |
| --- | --- | --- | --- | --- | --- | --- | --- | --- | --- |
| meta-analysis EAS | rs6006426 | 22 | 30669883 | G | A | 11,841/197,373 | 1.097 (1.066, 1.130) | $3.31 \times 10^{-10}$ | 0.079 |
| SCHS |  |  |  |  |  | 1,610/24,015 | 1.099 (1.018, 1.187) | 0.016 |  |
| Vietnamese |  |  |  |  |  | 1,598/1,267 | 1.169 (1.032, 1.324) | 0.014 |  |
| Han Chinese | | | | | | 833/1,220 | 1.282 (1.120, 1.468) | $3.16 \times 10^{-4}$ | |
| Biobank Japan | | | | | | 7,800/170,871 | 1.082 (1.046, 1.119) | $3.69 \times 10^{-6}$ | |
| meta-analysis EAS+EUR | | | | | | 12,736/673,864 | 1.098 (1.068, 1.129) | $4.33 \times 10^{-11}$ | 0.148 |
| UKB+FinnGen |  |  |  |  |  | 895/476,491 | 1.101 (1.002, 1.211) | 0.046 |  |

OR: Odds ratio; CI: confidence interval; EAS: East Asian; EUR: European; SCHS: Singapore Chinese Health Study; UKB: UK Biobank.

We assessed the transferability of the above association using summary statistics from European populations (16). A significant association was observed [OR (95%Cl) =1.101(1.002, 1.211), *P*=0.046, Table 1], and the overall meta-analysis was further strengthened after combining data from EAS and EUR [OR (95%Cl) =1.098(1.068, 1.129), *P_meta_*=4.33×10^-11^, Table 1].

### Functional mapping and annotations

rs6006426 and those in LD (r^2^ > 0.2) with it, are noted in the GWAS catalog (8) for their associations with a range of traits (Supplemental Table 2), yet there have been no associations reported with TB at genome-wide level of significance.

Regional information for rs6006426 displayed using LocusZoom (http://locuszoom.org/) revealed that this locus is situated in a gene-rich area that encompasses multiple genes within ±400kb of the lead SNP (Figure 1c). To prioritise genes potentially responsible for this association, we conducted gene-based test using MAGMA (v1.08), which uses a multiple regression approach to properly incorporate LD between markers and to detect multi-marker effects (21). This was implemented in FUMA GWAS (Functional Mapping and Annotation of Genome Wide Association Studies) (22) using the East Asian meta-analysis data. In addition to the Major Histocompatibility Complex (MHC) region on chromosome 6, *OSM* was significantly associated with PTB (ZSTAT=5.013; *P*=2.68×10^-7^; *P_adj_*=0.005) (Figure 2, Supplemental Table 3).

**Figure 2.**
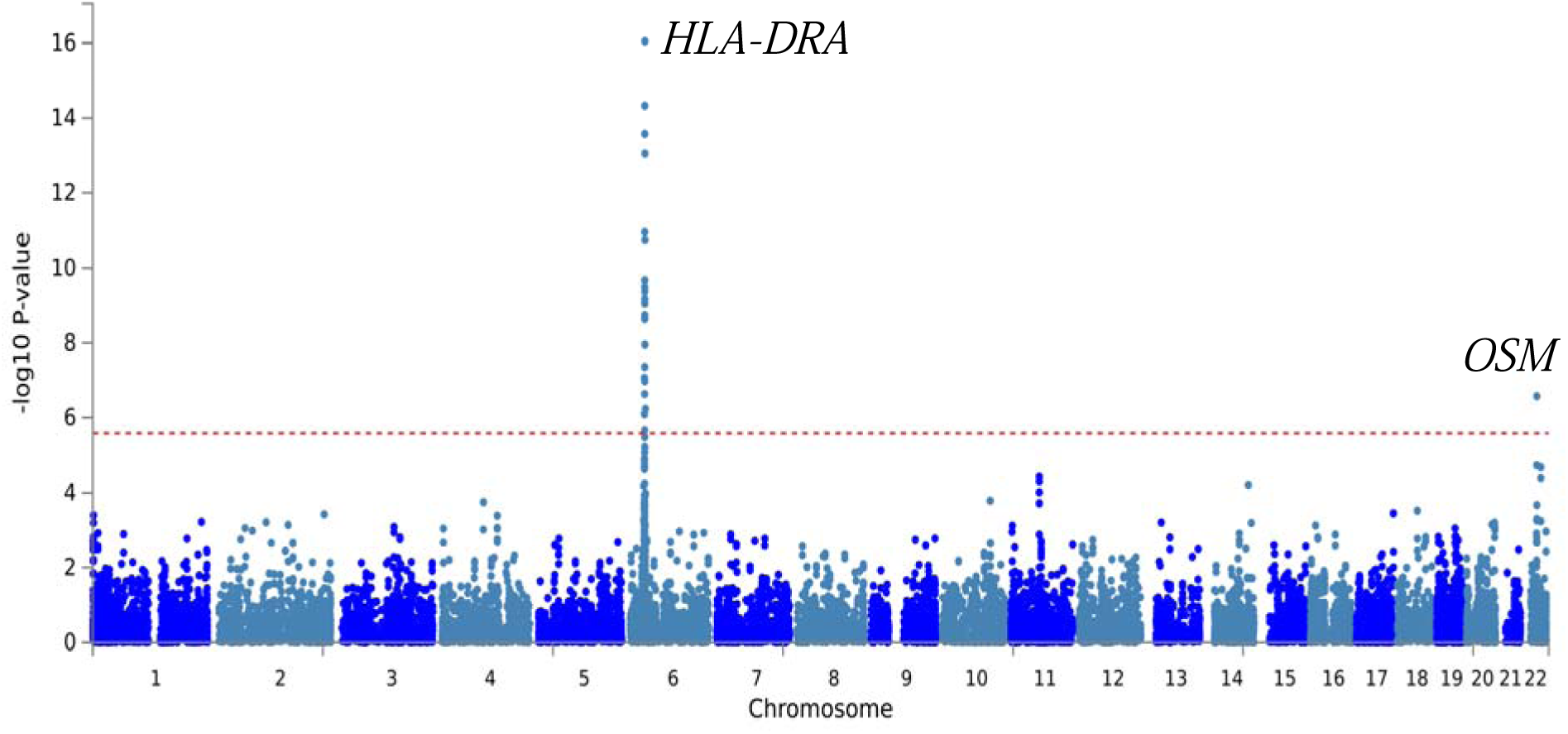
Manhattan plot of the gene-based test as computed by MAGMA using results from meta-analysis (11,841 cases/197,373 controls) as input. Input SNPs were mapped to 18,840 protein coding genes. Genome wide significance (red dashed line in the plot) was defined at P = 0.050/18,840 = 2.65×10^-6^.

We examined published TB transcriptomic and proteomic data to explore the potential functional link of candidate genes (within ±400kb of rs6006426) with TB (23–26). In African and mixed populations, *OSM*, *TCN2*, and *THOC5* were differentially expressed in various TB phenotypic comparisons and *LIF* had higher protein abundance in those who progressed to active TB (Supplemental Table 4) (23–26). To explore function of these genes in Asian populations, we assessed differential gene expression in monocytes of active PTB and healthy controls, from a previously published (27) mixed ancestry population of Singaporean Chinese, Malays, and Indians (20 PTB cases and 19 controls). *SF3A1*, *ASCC, LIF* and *MTMR3* showed differential expression, with the most significant, *SF3A1*, being downregulated in monocytes from PTB patients (Supplemental Figure 2). Together, these analyses highlighted seven candidate genes with potential functional relevance to TB pathogenesis. We then queried eQTL in immune cells from the eQTL Catalogue. rs6006426 significantly influences *SF3A1* expression in various immune cell regardless of exposure to viral, bacterial, or molecular stimuli (*P* ranging from 0.003 to 6.17×10^-18^, Supplemental Table 5) (28–30). The PTB risk increasing allele (allele A) was associated with reduced *SF3A1* expression. rs6006426 was not significantly associated with *OSM* expression in naive monocytes in most studies, however following stimulation with lipopolysaccharide (LPS) for 2 hours, the PTB risk allele significantly decreased *OSM* expression (β=-0.305, *P*=5.57×10^-4^, Supplemental Table 5) (30). The difference of *OSM* expression level among genotypes decreased with time and became less significant after 24 hours (β=-0.089, *P*=0.016, Supplemental Table 5) (30). From these data, *SF3A1* and *OSM* emerged as plausible candidate genes.

### Function of *sf3a1* and *osm* in the zebrafish-*M.marinum* infection model

Zebrafish are a valuable model for studying mycobacterial pathogenesis and treatment. Infection with *M. marinum*, the closest relative of the *Mtb* complex, mirrors many key aspects of human tuberculosis (31). *SF3A1* and *OSM* were individually knocked down in zebrafish embryos and the infection burden of *M. marinum* was assessed. Using standard dose CRISPR-Cas9 injections to knock down *sf3a1*, we observed a higher *M. marinum* burden in the crispants compared to the scramble control (Supplementary Figure 3a). However, the survival rate of these crispants was low, and they showed noticeable embryo morphological abnormalities (Supplementary Figure 3b). Upon reducing the CRISPR-Cas9 injection dose to a level that did not elicit morphological abnormalities, the *M. marinum* burden became comparable (Supplementary Figure 3c). Therefore, we hypothesize that the initial infection phenotype was likely due to non-specific developmental defects consistent with the early broad expression pattern of *sf3a1* observed during development (32). Targeting of *osm* by CRISPR-Cas9 mutagenesis was well tolerated by zebrafish embryos (Figure 3a), and we observed a decreased burden of *M.marinum* in the *osm* knockdown crispants (Figure 3b and 3c), suggesting a protective role for *osm* suppression during infection.

**Figure 3.**
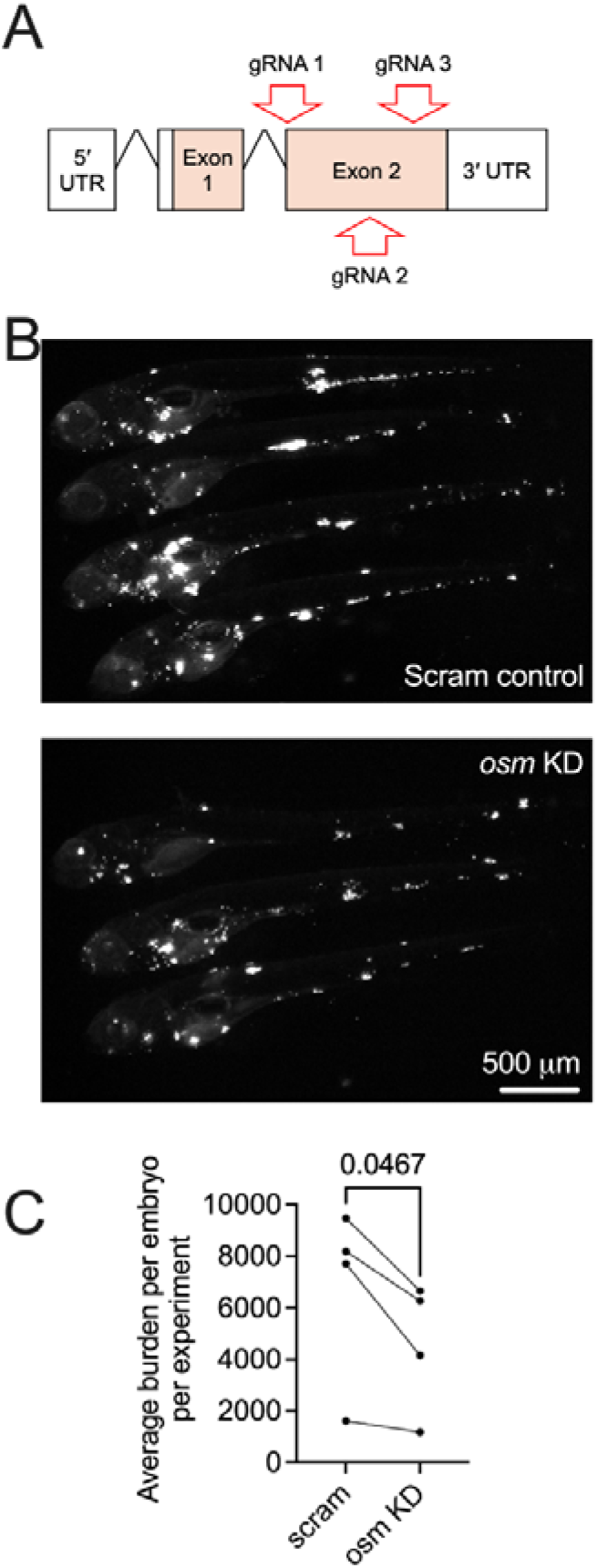
Functional analysis of *osm* in the zebrafish-*M. marinum* infection model. **a.** Schematic of the zebrafish *osm* genomic locus with CRISPR-Cas9 gRNA target sites annotated; **b.** Representative images of fluorescent *M. marinum* in *osm* knockdown zebrafish embryos; **c.** Comparison of average *M. marinum* infection burden in *osm* knockdown zebrafish embryos, n=4 experiments.

## Discussion

Genetic, environmental and socio-economic factors significantly contribute to TB susceptibility (3, 4). Our comprehensive GWAS with the largest sample size to date in East Asians identified a novel PTB susceptibility locus. The consistent significant association across 5 independent datasets has been rarely observed in prior TB GWAS. We identified *OSM* and *SF3A1* as potential candidate genes through functional mapping and annotation, neither of which has been reported in any genome-wide scaled analyses before. We showed *SF3A1* expression to be downregulated in monocytes from Singaporean PTB patients, and knocking down *osm* in a zebrafish model decreased the burden of *M. marinum* infection.

Previous GWAS have explored the genetic predisposition to TB in diverse populations (10–16) . These studies have yielded inconsistent results, which have been rarely replicable between studies. For instance, GWAS conducted in Ghanaian and Gambian identified 18q11.2 and 11p13 (10, 11), but only the 11p13 locus was replicated in Russia (12) and the South African study (17). Additionally, the *ASAP1* gene was reported in a Russian cohort (12), but was not replicated in an Icelandic population (13). These inconsistencies could stem from various factors, such as differing LD structures in distinct populations or phenotypic heterogeneity in case or control ascertainment. As *Mtb* lineages display substantial geographic variation and have been suggested to have co-evolved with human populations (33), pathogen genomic variation might also underlie some of the challenges in replicating across populations (34). These factors contribute to the increased complexity of conducting successful TB GWAS, in addition to performing GWAS in under-studied ethnicities that lack extensive reference genome panels. These findings underscore the importance of exercising caution when extrapolating GWAS results to different populations, highlighting the challenges of translating genetic associations across diverse ethnic groups. Conversely, our consistently replicated results show that combining datasets from genetically similar backgrounds enhances analytical power and facilitates discovery of novel TB loci.

The novel locus at 22q12.2 was supported by both single-variant and gene-based analyses. Multiple lines of evidence support *SF3A1* and *OSM* as the genes driving the association. *SF3A1* encodes a component of the splicing factor 3a (SF3A) protein complex, involved in pre-mRNA splicing as part of the spliceosome (35). A previous study reported that the immune system responds to infections through different mechanisms, one of which is alternative splicing (36). Dysregulation of, or mutations in, *SF3A1* may disrupt its role in pre-mRNA splicing, potentially affecting the immune response to bacterial infections. Inhibition of SF3A1 or SF3B1 increased production of a short form of MyD88 mRNA (MyD88_S_), a negative regulator of innate immunity through Toll-like receptor (TLR) signalling (37). MyD88_S_ level is critical in determining the production of inflammatory cytokines in murine macrophages (38). SF3A and SF3B mRNA splicing complexes function together in TLR signalling to regulate the production of MyD88_S_, and thereby control the extent of innate immune activation (37, 38). Considering that innate immune cells are the initial line of defence against *Mtb* (39) and that *Mtb* can potentially be eradicated by the innate immune system before the onset of an adaptive immune response (40), the modulation of *SF3A1* expression is particularly significant. Our findings suggest that the A allele of rs6006426 may exhibit reduced *SF3A1* expression, potentially leading to higher MyD88_S_ mRNA level, which could limit the activation of the innate immune system, rendering those exposed and infected more susceptible to progression to active TB disease. A previous candidate gene study in the Han Chinese established a connection between *SF3A1* and TB (41). Our sentinel SNP showed weak LD with the previously reported variants, rs2074733 (r^2^: 0.233 to 0.336) and rs10376 (r^2^: 0.056 to 0.099, EAS in 1000 Genomes Project East Asian). While rs2074733 was associated with PTB in 3 of our datasets, the direction of effect was inconsistent, potentially due to variations in case group composition. No association with rs10376 was found in our analysis (Supplemental Table 7). The functional impact of *sf3a1* could not be evaluated in the *M.marinum*-zebrafish infection model due to the non-specific developmental defects, and the potential essentiality of this gene. We propose further follow up studies are necessary using *in vitro* or monocyte/macrophage lineage-specific *in vivo* approaches to circumvent the role of *sf3a1* in embryonic development and reveal any TB-related phenotypes.

*OSM* is a secreted cytokine and growth regulator. eQTL data indicated that rs6006426 modulates *OSM* expression in monocytes after treatment with LPS, with the PTB risk allele significantly decreasing *OSM* expression. As LPS is a component of gram-negative bacterial cell walls that triggers TLR4 signalling (42), the eQTL effects might differ following stimulation of monocytes by *Mtb* ligands, or *Mtb* infection. A bioinformatics-based study reported *OSM* to be one of five upregulated genes predicting progression of latent tuberculosis infection (LTBI) to active TB in African and UK cohorts (43). A mechanistic study using cell culture and mouse models found *Mtb*-induced OSM produced by leukocytes drives expression of matrix metalloproteinases MMP-1 and MMP-3, and suppression of matrix metalloproteinase inhibitors, by pulmonary fibroblasts (44). OSM may therefore drive tissue destruction and facilitate dissemination of active TB and exacerbation of latent/subclinical TB. Further in vivo studies are required to decipher the TB context-dependent mechanism of allele-specific OSM-induced susceptibility and to link monocyte/macrophage production of OSM to granuloma matrix remodelling. The region identified in this large-scale meta-analysis may represent a shared regulatory area for both *OSM* and *SF3A1*, which together influence PTB risk. This mechanism might work similarly to how JAZF1 and TSPAN8 interact in diabetes (45). Further functional experiments are necessary to uncover the exact mechanism involved.

The primary strength of our study lies in the integrative meta-analysis of multiple East Asian populations, significantly augmenting the sample size and enhancing statistical power. However, the use of the general population as control subjects is a limitation. We combined individuals with a range of TB exposure statuses, including those never exposed to *Mtb*, those exposed but not infected, and those who have been exposed and *Mtb* infected but remain asymptomatic. This heterogeneity within the control group potentially leads to an underestimation of the true effect size of the association but is unlikely to result in false positive findings.

In conclusion, our meta-analysis identified 22q12.2 as a novel susceptibility locus for PTB in both East Asian and European populations. Although multiple lines of functional evidence suggest *SF3A1* and *OSM* as plausible candidate genes, the actual causal gene has not yet been fully determined. Our findings reveal new avenues for research, and further studies are necessary to elucidate the underlying mechanisms.

## Materials and Methods

### Study samples

The Singapore Chinese participants were enrolled in the SCHS, a prospective population-based cohort study of 63,257 participants (27,959 men and 35,298 women) aged 45 to 74 recruited between April 1993 and December 1998 to investigate genetic and environmental determinants of cancer and other chronic diseases in Singapore (19). Biospecimens were collected from a random 3% subset starting in 1994 and later expanded to all consenting participants (around half of the cohort). PTB cases were ascertained via linkage with the National Tuberculosis Notification Registry. Diagnosis is predominantly driven by passive case detection, when patients present with symptoms, such as persistent cough, blood-stained sputum, fever, chills, and night sweats. Cases were diagnosed by positive sputum smear and confirmed by microbiological culture of *Mtb*. Notifications primarily originate from public hospitals and the Tuberculosis Control Unit, supplemented by electronic records from the two Mycobacterial laboratories in Singapore, ensuring comprehensive data capture in the National Tuberculosis Notification Registry.

Blood samples were collected from PTB patients from Ho Chi Minh City (HCMC), Vietnam recruited as part of a larger clinical study (46). Briefly, 2,091 newly diagnosed PTB patients attending Pham Ngoc Thach Hospital outpatients department or one of 8 District TB Units (DTUs) in HCMC were recruited between December 2008 and July 2011. Patients sampled for the genetics study were 18 years or older, HIV negative, had no previous history of TB treatment and were sputum smear positive. DNA was extracted using Qiagen Blood Midi kits (Qiagen) and 1,650 underwent whole genome genotyping at the Genome Institute of Singapore. Vietnamese Kinh population controls are otherwise healthy adults with primary angle closure glaucoma who have been previously described (47).

SCHS was approved by the Institutional Review Board (IRB) at the National University of Singapore. The study involving Vietnamese was approved by the IRBs of the Hospital for Tropical Diseases HCMC Vietnam, Pham Ngoc Thach Hospital for Tuberculosis and Lung Disease HCMC Vietnam, Health Services of HCMC Vietnam, the Oxford Tropical Research Ethics Committee, Oxford University UK and the University of Melbourne Human Research Ethics Committee, Melbourne Australia (ID 21973). Written informed consent was obtained from all study participants for both studies.

### Genotyping and Imputation

In SCHS, 27,308 DNA samples were whole genome genotyped using the Illumina Global Screening Array (GSA). An additional 2,161 independent subjects from the SCHS CAD- nested case-control study were genotyped on Illumina HumanOmniZhonghua Bead Chip. Comprehensive information regarding genotyping and quality control (QC) protocols has been previously published (48). After QC, SNP alleles were standardized to the forward strand and mapped to the hg38 reference genome. Minimac4 (version 1.0.0) (49) was employed to impute additional autosomal SNPs using a local population-specific reference panels compromised of 9,770 whole-genome sequences of local Singaporean population samples obtained from the SG10K initiative (SG10K Health) (50) on the Research Assets Provisioning and Tracking Online Repository (RAPTOR) (51).

The Vietnamese samples were genotyped using either the Infinium OmniExpress-24 or OmniExpress-12 array. 1,650 PTB cases were combined with 1,357 Vietnamese kinh population controls. Samples with call-rate <95% or extreme heterozygosity (beyond ±3 standard deviations, N=47) were excluded. Identity-by-state (IBS) analyses identified first and second-degree related samples, with lower call-rate sample from each detected pair removed (N=83). PCA with 1000 Genomes Projects reference populations and within the Vietnamese samples identified and removed 12 ethnicity outliers. For SNP QC (Supplemental Table 6), monomorphic or rare SNPs (MAF<1.0%), those with call-rates <95.0% (N=112,360), sex chromosome SNPs, SNPs shown different missingness (N=7,088), and those displaying gross Hardy–Weinberg equilibrium (HWE) deviation (*P* < 10^-6^, N=13,601) were removed. SNPs were coded to the forward strand and mapped to hg19. IMPUTE v2 (52) was used to mutually impute variants with cosmopolitan 1000 Genomes haplotypes as reference panel (Phase 3) (20). SNPs with impute information score < 0.6, MAF < 1.0% or non-biallelic SNPs were excluded from subsequent analyses.

### Statistical analysis

Sex was not considered as a biological variable in our study. Single-variant association tests were performed using genotype dosage data with an R package SAIGE (v1.3.0) (53), adjusted for the first three principal components to correct for the population stratification. SAIGE accounts for sample relatedness and manages case-control imbalance of binary traits. Genome-wide significance threshold was set at 5×10^-8^. To assess potential inflation in the study results, the genomic inflation factor (λ) was calculated. Publicly available association summary statistics were downloaded from Biobank Japan (16) (https://pheweb.jp/pheno/PTB) and a Han Chinese dataset (15) (https://doi.org/10.6084/m9.figshare.7006310). Summary statistics from European populations published in the PTB GWAS from Biobank Japan were also downloaded (16). An inverse variance-weighted meta-analysis was employed using META (v1.7) (54), assuming a fixed effects model. Heterogeneity among the combined data was evaluated using Cochran’s Q (55) and a *P*-value (*P*_heterogeneity_) < 0.05 was determined to be significantly heterogeneous.

### Functional mapping and annotation

Lead SNPs discovered were functionally annotated using the SNP2GENE function in FUMA (v1.6.1) (22). SNPs in LD (r^2^ > 0.6 in 1000G ASN panel) with sentinel SNPs were identified. Genes located within a 10 kb region flanking each lead SNP were mapped as regional genes at the locus of interest. FUMA performs MAGMA gene analysis (v1.08) (21) using the default SNP-wide mean model and 1000G EAS population as a reference panel. Previously published research on TB transcriptomics and proteomics was explored to identify TB candidate genes (23–26). Differentially expressed genes in monocytes between active PTB cases and controls (GSE126614) were also analysed in a mixed ethnicity population (27). Cell-type specific eQTL data from eQTL Catalogue (56) was queried; the significance of eQTLs was assessed based on the number of potential candidate genes under investigation (*P*=0.050/7=0.007).

### Knockdown of *sf3a1* and *osm* in zebrafish embryos and analysis of *M. marinum* infection burden

Four guide RNAs per gene target were prepared using a common scaffold oligo (AAAAGCACCGACTCGGTGCCACTTTTTCAAGTTGATAACGGACTAGCCTTATTTT AACTTGCTATTTCTAGCTCTAAAAC) and *in vitro* transcription (New England Biolabs) (57). Scramble control: TAATACGACTCACTATAGGCAGGCAAAGAATCCCTGCCGTTTTAGAGCTAGAAATAGC, TAATACGACTCACTATAGGTACAGTGGACCTCGGTGTCGTTTTAGAGCTAGAAATA GC, TAATACGACTCACTATAGGCTTCATACAATAGACGATGGTTTTAGAGCTAGAAATAGC, TAATACGACTCACTATAGGTCGTTTTGCAGTAGGATCGGTTTTAGAGCTAGAAATAGC; *sf3a1*: TAATACGACTCACTATAGGGTCGCAGTCATGCCGCCTGTTTTAGAGCTAGAAATAGC, TAATACGACTCACTATAGGACTGCCAGCTTCGTAGCCGTTTTAGAGCTAGAAATAGC, TAATACGACTCACTATAGGTATGGGTCGCTCGGGTTGGTTTTAGAGCTAGAAATAGC, TAATACGACTCACTATAGGAGGTGATCCATGAGACGGGTTTTAGAGCTAGAAATAGC; *osm*: TAATACGACTCACTATAGGTTACTTTGAGCTGAGAAGGTTTTAGAGCTAGAAATAGC, TAATACGACTCACTATAGGACACCAGGAGGATCTAGTGTTTTAGAGCTAGAAATAGC, TAATACGACTCACTATAGGGAGGACTCAATGACCGCGGTTTTAGAGCTAGAAATAGC. Injection solutions were contained 1 µl phenol red dye (Sigma), 2 µl 500 ng/µl pooled guides, and 2 µl 10 µM Cas9 (IDT DNA). Injections were performed into the yolk of 1-2 cells stage fertilized zebrafish eggs and embryos were reared at 28°C with the addition of PTU at 1 day post fertilization (dpf). Embryos were infected with ∼200 CFU fluorescent *M. marinum* at 2 dpf and imaged for bacterial burden quantification by fluorescent pixel count at 5 dpf (58). Bacterial burden was averaged for each experiment and compared across 4 replicates by paired *t*-test (Graphpad Prism).

## Data Availability

Summary statistics for PTB GWAS in Singaporean Chinese and Vietnamese will be available in figshare on publication.

## Acknowledgements

We wish to thank the individuals who participated in our studies. We acknowledge the clinical staff who recruited patients in the Vietnam study from the following District TB Units (DTUs) in Ho Chi Minh City: Districts 1, 4, 5, 6 and 8, Tan Binh, Binh Thanh and Phu Nhuan; and also our colleagues from Pham Ngoc Thach Hospital for Tuberculosis and Lung Disease, particularly Nguyen Ngoc Lan and Nguyen Huu Lan and from Oxford University Clinical Research Unit, Hoang Thanh Hai and Vu Thi Ngoc Ha. We thank Siew-Hong Low of the National University of Singapore for supervising the fieldwork in the Singapore Chinese Health Study.

## Author contributions

X.C., R.D, CCK, S.J.D were responsible for conceptualization and X.C., R.D, S.J.D wrote the manuscript. PVKT, DTMH, NTTT, NTQN, MC and S.J.D were responsible for the clinical study in Vietnam (including patient diagnosis, recruitment and sampling). CBEC, C-KH, J- MY, W-PK, RD, CCK, AS were responsible for data collection and data generation that contributed to this manuscript from the Singapore Chinese Health Study and Singapore TB study. ZL, CCK were responsible for all data generation and QC for the Vietnamese datasets. DW, ASBMS and SHO were responsible for all data generation and analysis involving the *M.marinum* zebrafish model. All analysis was performed by XC and RD. S.J.D, CCK, MC, YYK was responsible for funding acquisition for the Vietnamese study. All authors reviewed and edited the manuscript.

## References

1. World Health Organization. Global tuberculosis report 2024 (World Health Organization. 29 October 2024)

2. García JI, Allué-Guardia A, Tampi RP, Restrepo BI, Torrelles JB. New Developments and Insights in the Improvement of Mycobacterium tuberculosis Vaccines and Diagnostics Within the End TB Strategy. Current Epidemiology Reports. 2021;8(2):33–45.

3. Narasimhan P, Wood J, Macintyre CR, Mathai D. Risk factors for tuberculosis. Pulm Med. 2013;2013:828939.

4. Qu H-Q, Fisher-Hoch SP, McCormick JB. Knowledge gaining by human genetic studies on tuberculosis susceptibility. Journal of Human Genetics. 2011;56(3):177–82.

5. Apt A, Kramnik I. Man and mouse TB: contradictions and solutions. Tuberculosis (Edinb). 2009;89(3):195–8.

6. Comstock GW. Tuberculosis in twins: a re-analysis of the Prophit survey. Am Rev Respir Dis. 1978;117(4):621–4.

7. Abel L, El-Baghdadi J, Bousfiha AA, Casanova JL, Schurr E. Human genetics of tuberculosis: a long and winding road. Philos Trans R Soc Lond B Biol Sci. 2014;369(1645):20130428.

8. Sollis E, Mosaku A, Abid A, Buniello A, Cerezo M, Gil L, et al. The NHGRI-EBI GWAS Catalog: knowledgebase and deposition resource. Nucleic Acids Res. 2023;51(D1):D977–d85.

9. Mills MC, Rahal C. The GWAS Diversity Monitor tracks diversity by disease in real time. Nature Genetics. 2020;52(3):242–3.

10. Thye T, Vannberg FO, Wong SH, Owusu-Dabo E, Osei I, Gyapong J, et al. Genome-wide association analyses identifies a susceptibility locus for tuberculosis on chromosome 18q11.2. Nat Genet. 2010;42(9):739–41.

11. Thye T, Owusu-Dabo E, Vannberg FO, van Crevel R, Curtis J, Sahiratmadja E, et al. Common variants at 11p13 are associated with susceptibility to tuberculosis. Nat Genet. 2012;44(3):257–9.

12. Curtis J, Luo Y, Zenner HL, Cuchet-Lourenço D, Wu C, Lo K, et al. Susceptibility to tuberculosis is associated with variants in the ASAP1 gene encoding a regulator of dendritic cell migration. Nature Genetics. 2015;47(5):523–7.

13. Sveinbjornsson G, Gudbjartsson DF, Halldorsson BV, Kristinsson KG, Gottfredsson M, Barrett JC, et al. HLA class II sequence variants influence tuberculosis risk in populations of European ancestry. Nat Genet. 2016;48(3):318–22.

14. Qi H, Zhang Y-B, Sun L, Chen C, Xu B, Xu F, et al. Discovery of susceptibility loci associated with tuberculosis in Han Chinese. Human Molecular Genetics. 2017;26(23):4752–63.

15. Zheng R, Li Z, He F, Liu H, Chen J, Chen J, et al. Genome-wide association study identifies two risk loci for tuberculosis in Han Chinese. Nature Communications. 2018;9(1):4072.

16. Sakaue S, Kanai M, Tanigawa Y, Karjalainen J, Kurki M, Koshiba S, et al. A cross-population atlas of genetic associations for 220 human phenotypes. Nature Genetics. 2021;53(10):1415–24.

17. Chimusa ER, Zaitlen N, Daya M, Möller M, van Helden PD, Mulder NJ, et al. Genome-wide association study of ancestry-specific TB risk in the South African Coloured population. Hum Mol Genet. 2014;23(3):796–809.

18. Kerimov N, Hayhurst JD, Peikova K, Manning JR, Walter P, Kolberg L, et al. A compendium of uniformly processed human gene expression and splicing quantitative trait loci. Nature Genetics. 2021;53(9):1290–9.

19. Hankin JH, Stram DO, Arakawa K, Park S, Low S-H, Lee H-P, et al. Singapore Chinese Health Study: development, validation, and calibration of the quantitative food frequency questionnaire. Nutrition and cancer. 2001;39(2):187–95.

20. Auton A, Abecasis GR, Altshuler DM, Durbin RM, Abecasis GR, Bentley DR, et al. A global reference for human genetic variation. Nature. 2015;526(7571):68-74.

21. de Leeuw CA, Mooij JM, Heskes T, Posthuma D. MAGMA: Generalized Gene-Set Analysis of GWAS Data. PLOS Computational Biology. 2015;11(4):e1004219.

22. Watanabe K, Taskesen E, Van Bochoven A, Posthuma D. Functional mapping and annotation of genetic associations with FUMA. Nature communications. 2017;8(1):1826.

23. Berry MPR, Graham CM, McNab FW, Xu Z, Bloch SAA, Oni T, et al. An interferon-inducible neutrophil-driven blood transcriptional signature in human tuberculosis. Nature. 2010;466(7309):973-7.

24. Kaforou M, Wright VJ, Oni T, French N, Anderson ST, Bangani N, et al. Detection of tuberculosis in HIV-infected and -uninfected African adults using whole blood RNA expression signatures: a case-control study. PLoS Med. 2013;10(10):e1001538.

25. Penn-Nicholson A, Hraha T, Thompson EG, Sterling D, Mbandi SK, Wall KM, et al. Discovery and validation of a prognostic proteomic signature for tuberculosis progression: A prospective cohort study. PLoS Med. 2019;16(4):e1002781.

26. Kroon EE, Correa-Macedo W, Evans R, Seeger A, Engelbrecht L, Kriel JA, et al. Neutrophil extracellular trap formation and gene programs distinguish TST/IGRA sensitization outcomes among Mycobacterium tuberculosis exposed persons living with HIV. PLoS Genet. 2023;19(8):e1010888.

27. Del Rosario RCH, Poschmann J, Lim C, Cheng CY, Kumar P, Riou C, et al. Histone acetylome-wide associations in immune cells from individuals with active Mycobacterium tuberculosis infection. Nat Microbiol. 2022;7(2):312–26.

28. Momozawa Y, Dmitrieva J, Théâtre E, Deffontaine V, Rahmouni S, Charloteaux B, et al. IBD risk loci are enriched in multigenic regulatory modules encompassing putative causative genes. Nature Communications. 2018;9(1):2427.

29. Fairfax BP, Makino S, Radhakrishnan J, Plant K, Leslie S, Dilthey A, et al. Genetics of gene expression in primary immune cells identifies cell type–specific master regulators and roles of HLA alleles. Nature Genetics. 2012;44(5):502–10.

30. Fairfax BP, Humburg P, Makino S, Naranbhai V, Wong D, Lau E, et al. Innate immune activity conditions the effect of regulatory variants upon monocyte gene expression. Science. 2014;343(6175):1246949.

31. Cronan MR, Tobin DM. Fit for consumption: zebrafish as a model for tuberculosis. Dis Model Mech. 2014;7(7):777–84.

32. Sur A, Wang Y, Capar P, Margolin G, Prochaska MK, Farrell JA. Single-cell analysis of shared signatures and transcriptional diversity during zebrafish development. Dev Cell. 2023;58(24):3028–47.e12.

33. Silcocks M, Dunstan SJ. Parallel signatures of Mycobacterium tuberculosis and human Y-chromosome phylogeography support the Two Layer model of East Asian population history. Commun Biol. 2023;6(1):1037.

34. Gagneux S, DeRiemer K, Van T, Kato-Maeda M, de Jong BC, Narayanan S, et al. Variable host-pathogen compatibility in Mycobacterium tuberculosis. Proc Natl Acad Sci U S A. 2006;103(8):2869–73.

35. Visconte V, M ON, H JR. Mutations in Splicing Factor Genes in Myeloid Malignancies: Significance and Impact on Clinical Features. Cancers (Basel). 2019;11(12).

36. Hong W, Yang H, Wang X, Shi J, Zhang J, Xie J. The Role of mRNA Alternative Splicing in Macrophages Infected with Mycobacterium tuberculosis: A Field Needing to Be Discovered. Molecules. 2024;29(8).

37. De Arras L, Alper S. Limiting of the innate immune response by SF3A-dependent control of MyD88 alternative mRNA splicing. PLoS Genet. 2013;9(10):e1003855.

38. O’Connor BP, Danhorn T, De Arras L, Flatley BR, Marcus RA, Farias-Hesson E, et al. Regulation of Toll-like Receptor Signaling by the SF3a mRNA Splicing Complex. PLOS Genetics. 2015;11(2):e1004932.

39. Ravesloot-Chávez MM, Dis EV, Stanley SA. The Innate Immune Response to Mycobacterium tuberculosis Infection. Annual Review of Immunology. 2021;39(1):611–37.

40. Lerner TR, Borel S, Gutierrez MG. The innate immune response in human tuberculosis. Cell Microbiol. 2015;17(9):1277–85.

41. Zhang J, Wang M-G, Liu Q-x, He J-Q. Association between a single nucleotide polymorphism of the SF3A1 gene and tuberculosis in a Chinese Han population: a case[control study21 November 2022, PREPRINT (Version 1) available at Research Square [10.21203/rs.3.rs-2252919/v1].

42. Mazgaeen L, Gurung P. Recent Advances in Lipopolysaccharide Recognition Systems. Int J Mol Sci. 2020;21(2).

43. Ma S, Peng P, Duan Z, Fan Y, Li X. Predicting the Progress of Tuberculosis by Inflammatory Response-Related Genes Based on Multiple Machine Learning Comprehensive Analysis. J Immunol Res. 2023;2023:7829286.

44. O’Kane CM, Elkington PT, Friedland JS. Monocyte-dependent oncostatin M and TNF-alpha synergize to stimulate unopposed matrix metalloproteinase-1/3 secretion from human lung fibroblasts in tuberculosis. Eur J Immunol. 2008;38(5):1321–30.

45. Grarup N, Andersen G, Krarup NT, Albrechtsen A, Schmitz O, Jørgensen T, et al. Association testing of novel type 2 diabetes risk alleles in the JAZF1, CDC123/CAMK1D, TSPAN8, THADA, ADAMTS9, and NOTCH2 loci with insulin release, insulin sensitivity, and obesity in a population-based sample of 4,516 glucose-tolerant middle-aged Danes. Diabetes. 2008;57(9):2534–40.

46. Thai PVK, Ha DTM, Hanh NT, Day J, Dunstan S, Nhu NTQ, et al. Bacterial risk factors for treatment failure and relapse among patients with isoniazid resistant tuberculosis. BMC Infectious Diseases. 2018;18(1):112.

47. Khor CC, Do T, Jia H, Nakano M, George R, Abu-Amero K, et al. Genome-wide association study identifies five new susceptibility loci for primary angle closure glaucoma. Nat Genet. 2016;48(5):556–62.

48. Chang X, Gurung RL, Wang L, Jin A, Li Z, Wang R, et al. Low frequency variants associated with leukocyte telomere length in the Singapore Chinese population. Communications Biology. 2021;4(1):519.

49. Das S, Forer L, Schönherr S, Sidore C, Locke AE, Kwong A, et al. Next-generation genotype imputation service and methods. Nature Genetics. 2016;48(10):1284–7.

50. Wong E, Bertin N, Hebrard M, Tirado-Magallanes R, Bellis C, Lim WK, et al. The Singapore National Precision Medicine Strategy. Nat Genet. 2023;55(2):178–86.

51. Shih CC, Chen J, Lee AS, Bertin N, Hebrard M, Khor CC, et al. A five-safes approach to a secure and scalable genomics data repository. iScience. 2023;26(4):106546.

52. Marchini J, Howie B, Myers S, McVean G, Donnelly P. A new multipoint method for genome-wide association studies by imputation of genotypes. Nat Genet. 2007;39(7):906–13.

53. Zhou W, Nielsen JB, Fritsche LG, Dey R, Gabrielsen ME, Wolford BN, et al. Efficiently controlling for case-control imbalance and sample relatedness in large-scale genetic association studies. Nature Genetics. 2018;50(9):1335–41.

54. Liu JZ, Tozzi F, Waterworth DM, Pillai SG, Muglia P, Middleton L, et al. Meta-analysis and imputation refines the association of 15q25 with smoking quantity. Nat Genet. 2010;42(5):436–40.

55. Conover WJ. Practical nonparametric statistics: john wiley & sons; 1999.

56. Kerimov N, Tambets R, Hayhurst JD, Rahu I, Kolberg P, Raudvere U, et al. eQTL Catalogue 2023: New datasets, X chromosome QTLs, and improved detection and visualisation of transcript-level QTLs. PLOS Genetics. 2023;19(9):e1010932.

57. Wu RS, Lam, II, Clay H, Duong DN, Deo RC, Coughlin SR. A Rapid Method for Directed Gene Knockout for Screening in G0 Zebrafish. Dev Cell. 2018;46(1):112–25.e4.

58. Matty MA, Oehlers SH, Tobin DM. Live Imaging of Host-Pathogen Interactions in Zebrafish Larvae. Methods Mol Biol. 2016;1451:207–23.

