## Supplemental Figures for "Genome-wide association study reveals a novel tuberculosis susceptibility locus in multiple East Asian and European populations"

### Slide 1
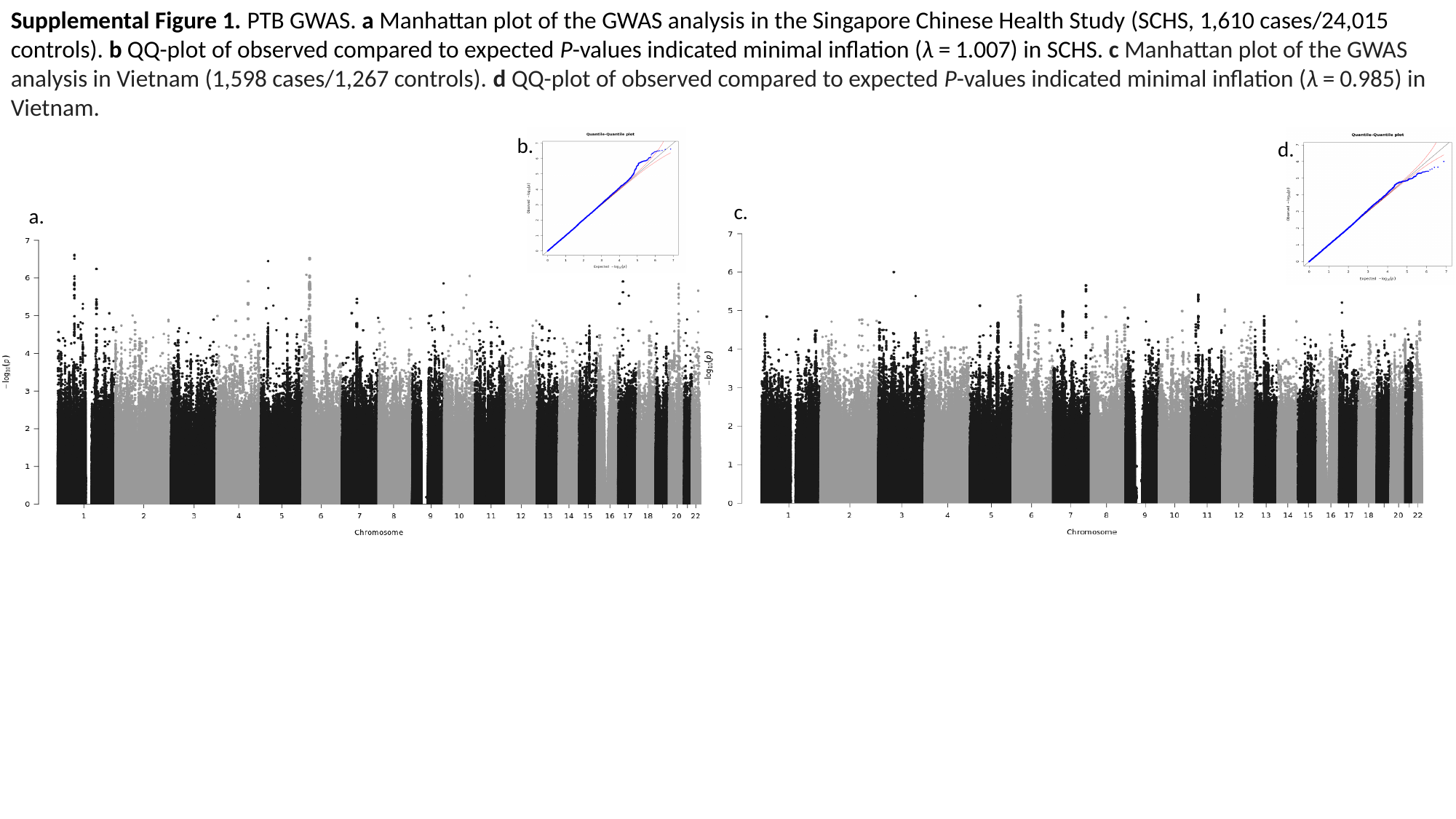

Supplemental Figure 1. PTB GWAS. a Manhattan plot of the GWAS analysis in the Singapore Chinese Health Study (SCHS, 1,610 cases/24,015 controls). b QQ-plot of observed compared to expected P-values indicated minimal inflation (λ = 1.007) in SCHS. c Manhattan plot of the GWAS analysis in Vietnam (1,598 cases/1,267 controls). d QQ-plot of observed compared to expected P-values indicated minimal inflation (λ = 0.985) in Vietnam.
b.
a.
d.
c.

### Slide 2
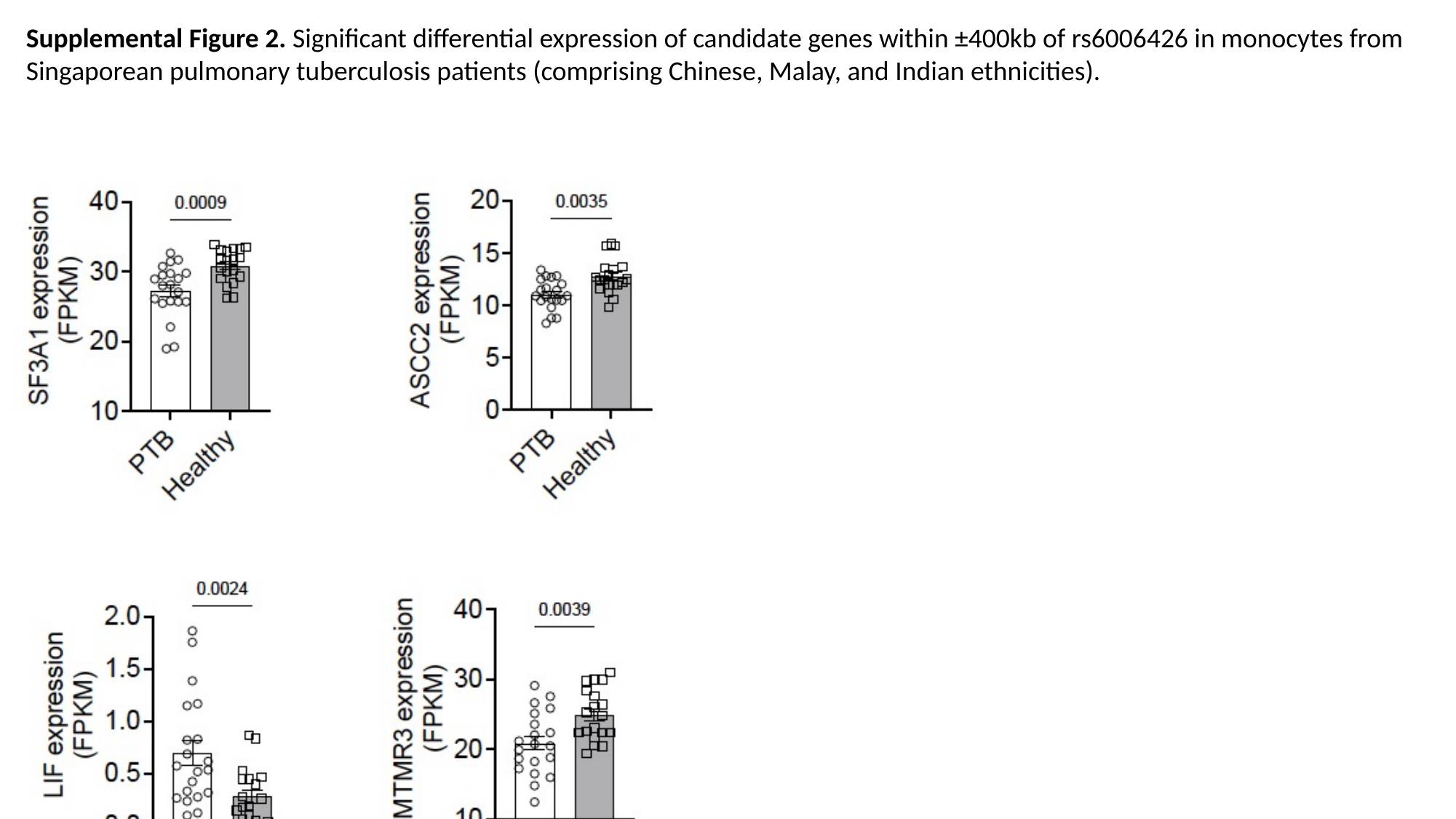

Supplemental Figure 2. Significant differential expression of candidate genes within ±400kb of rs6006426 in monocytes from Singaporean pulmonary tuberculosis patients (comprising Chinese, Malay, and Indian ethnicities).

### Slide 3
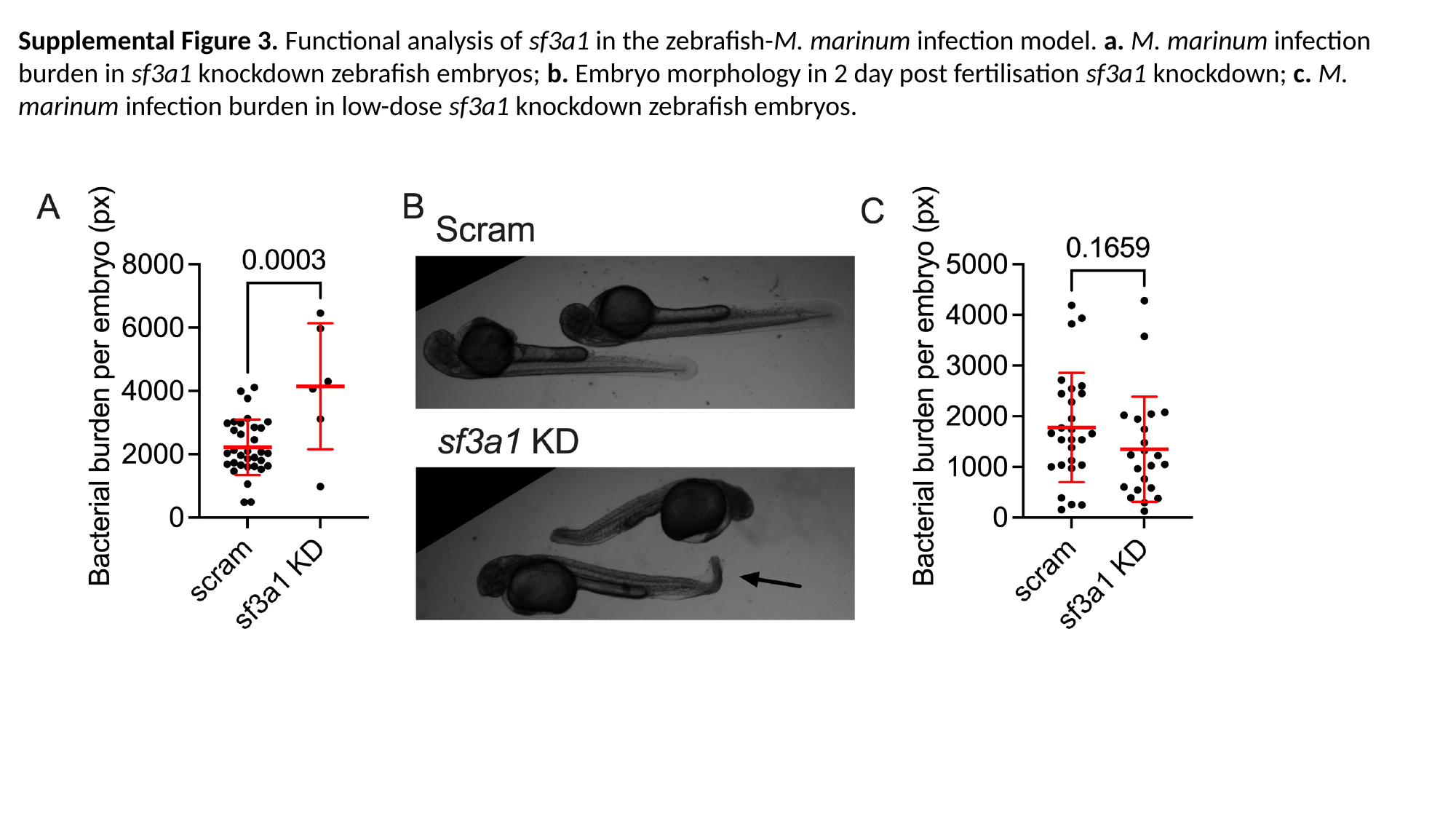

Supplemental Figure 3. Functional analysis of sf3a1 in the zebrafish-M. marinum infection model. a. M. marinum infection burden in sf3a1 knockdown zebrafish embryos; b. Embryo morphology in 2 day post fertilisation sf3a1 knockdown; c. M. marinum infection burden in low-dose sf3a1 knockdown zebrafish embryos.
